# Respiratory symptoms of Swiss people with Primary Ciliary Dyskinesia

**DOI:** 10.1101/2021.11.29.21266978

**Authors:** Myrofora Goutaki, Leonie Hüsler, Yin Ting Lam, Helena M. Koppe, Andreas Jung, Romain Lazor, Loretta Müller, Swiss PCD research group, Eva SL Pedersen, Claudia E. Kuehni

**Affiliations:** Institute of Social and Preventive Medicine, University of Bern, Bern, Switzerland; Division of Paediatric Respiratory Medicine and Allergology, Department of Paediatrics, Inselspital, Bern University Hospital, University of Bern, Bern, Switzerland; Paediatric Pulmonology, University Children’s Hospital Zurich, Zurich, Switzerland; Respiratory Medicine Department, Lausanne University Hospital, University of Lausanne, Lausanne, Switzerland; Department of BioMedical Research (DBMR), University of Bern, Switzerland

**Keywords:** Primary ciliary dyskinesia, epidemiology, orphan disease, respiratory symptoms, clinical variability

## Abstract

**Background:** Mostly derived from chart reviews, where symptoms are recorded in a nonstandardised manner, clinical data about primary ciliary dyskinesia (PCD) are inconsistent, which leads to missing and unreliable information. We assessed the prevalence and frequency of respiratory symptoms and studied differences by age and sex among an unselected population of Swiss people with PCD.

**Methods:** We sent a questionnaire that included items from the FOLLOW-PCD standardised questionnaire to all Swiss PCD registry participants.

**Results:** We received questionnaires from 74 out of 86 (86%) invited persons or their caregivers (age range: 3–73 years; median age: 23), including 68% adults (≥18 years) and 51% females. Among participants, 70 (94%) reported chronic nasal symptoms, most frequently runny nose (65%), blocked nose (55%), or anosmia (38%). Ear pain and hearing problems were reported by 58% of the participants. Almost all (99%) reported cough and sputum production. The most common chronic cough complications were gastroesophageal reflux (n=11; 15%), vomiting (n=8; 11%), and urinary incontinence (n=6; 8%). Only 9 participants (12%) reported frequent wheeze, which occurred mainly during infection or exercise, while 49 persons (66%) reported shortness of breath; 9% even at rest or during daily activities. Older patients reported more frequent nasal symptoms and shortness of breath. We found no difference by sex or ultrastructural ciliary defect.

**Conclusion:** This is the first study that describes patient-reported PCD symptoms. The consistent collection of standardised clinical data will allow us to better characterise the phenotypic variability of the disease and study disease course and prognosis.

**Take home message:** Detailed information about patient-reported PCD symptoms will help characterise the phenotypic variability of the disease and could inform the development of individualised treatment strategies for patients.

## INTRODUCTION

Primary ciliary dyskinesia (PCD) is a rare, genetic, multi-organ disease characterised by significant genetic and clinical variability [1,2]. Around 50 genes have been implicated in causing PCD, and diagnostic advances in recent years have been tremendous [3-6]. PCD seems to have substantial phenotypic variability, yet we lack data to confirm this [2,7,8]. Since the disease impairs mucociliary clearance in the airways, most persons affected by PCD develop upper and lower respiratory symptoms [9-11]. PCD can manifest within a few hours of birth; usually with unspecific neonatal rhinitis and cough or unexplained respiratory distress [10,12]. As patients grow, recurrent ear, sinus, and lung infections are common and many patients develop chronic rhinosinusitis, hearing impairment, and bronchiectasis [13-15].

Knowledge about symptom prevalence includes heterogeneous results derived mainly from hospital-based, single-centre studies [9]. Most of these studies described diagnostic data or other measurements, such as lung function test results, while only briefly reporting the prevalence of a few symptoms [9]. Symptom data were usually extracted from medical charts where they were not recorded in a standardised manner, leading to missing and unreliable information. Usually, study participants were either recruited from pulmonology departments for lower airway-focused studies or from ear, nose, and throat (ENT) clinics for studies including only patients with moderate to severe ENT symptoms [9]. No studies so far reported symptom frequency or severity because this information is rarely recorded in charts. Although PCD evolves with age and symptoms can change from childhood to adulthood, most studies only include children or young adults [8,16-18]. However, when studies included children and older adults, they rarely reported differences in symptom prevalence by age [9]. Similarly, data on phenotypic difference by sex is scarce, inconclusive, and relates mostly to lung function [19]. In this study, we assessed the prevalence and frequency of respiratory symptoms and how they differ by age and sex in an unselected population of Swiss people with PCD.

## METHODS

### Study design and study population

Our study wa s a cross-sectional questionnaire survey nested in the Swiss PCD registry (CH-PCD)[20]. CH-PCD is a patient registry (www.clinicaltrials.gov; identifier <u>NCT03606200</u>) that collects demographic characteristics, diagnostic test results, clinical data, and information about management of patients in Switzerland. It was established in 2013, and it provides data for national and international PCD monitoring and research [21-23]. Patients are identified by physicians, diagnostic facilities, or local support groups, such as the Swiss PCD support group, “Selbsthilfegruppe Kartagener Syndrom und Primäre Ciliäre Dyskinesie Deutschschweiz”. CH-PCD was approved by the Cantonal Ethics Committee of Bern in 2015 (KEK-BE: 060/2015). We obtained written informed consent from either people aged 14 years and older or from parents of participants younger than age 14. We followed the Strengthening the Reporting of Observational Studies in Epidemiology (STROBE) reporting recommendations [24].

All participants registered in CH-PCD who read German or French, signed an informed consent, and had a valid postal address were invited to complete the questionnaire. We excluded one patient who spoke only Italian and four patients with invalid postal addresses. We sent the questionnaire by post to German-speaking participants in February 2020 and to French-speaking participants in July 2020. No later than three weeks after the original questionnaire mailing, we sent a reminder letter to those who had not yet returned the questionnaire. Five newly registered CH-PCD participants who enrolled during the study period received the questionnaire at the PCD outpatient clinic in Bern during their follow-up visit.

### Questionnaire

We used an adapted version of the FOLLOW-PCD questionnaire (version 1.0), which asks about PCD-specific symptoms and health behaviours [25]. The questionnaire is part of the FOLLOW-PCD form for standardised4 clinical follow-up for PCD patients developed by an international, multidisciplinary group of PCD experts [25]. The first part of the questionnaire collects detailed information about the frequency and characteristics of upper and lower respiratory and general symptoms during the past three months. The second part focuses on environmental exposures and health-related behaviours during the past year. Specifically, it collects information about school or work attendance, physical activity, active and passive smoking, nutrition, and environmental exposures. There are three versions of the questionnaire designed for different age groups: a) a parent/caretaker questionnaire for children aged 0-13 years, b) an adolescent version for those aged 14–17 years, and c) an adult questionnaire for participants aged ≥18 years.

Our survey included all FOLLOW-PCD questions and additional ones developed for this study. We added questions about extrapulmonary manifestations of PCD (*e.g*. laterality defects, congenital heart defects) and triggers for wheezing [26,27]. We also added a section on physiotherapy, as well as more detailed questions about smoking and nutrition [28-30]. These topics are difficult to extract information about from medical charts since they are often inconsistently recorded. The Swiss PCD symptom survey was an excellent opportunity to assess this information directly from participants. We piloted the questionnaire among members of the Swiss PCD support group to ensure that the wording and content were well understood. The FOLLOW-PCD questionnaire was available in German and French. The additional questions were originally developed in German then translated into French by a native French speaker with knowledge about PCD.

### Respiratory symptoms and demographic information

The questionnaire inquired about upper respiratory symptoms (*e.g*. nasal symptoms, snoring, ear pain, hearing problems) and lower respiratory symptoms (*e.g*. cough, sputum production, wheezing, shortness of breath, chest pain) during the past 3 months. Questions about symptom frequency included five possible response categories: daily, often, sometimes, rarely, and never. We defined symptoms reported often or daily as frequent. For some symptoms, we asked about attributes such as persistence, diurnal variability, possible triggers, and quantity and colour of sputum. For adult patients, we assessed dyspnea by score using the modified Medical Research Council (mMRC) dyspnea scale. Sex, area of residence, and diagnostic tests were retrieved from CH-PCD. We calculated age at survey from the questionnaire completion date.

### Diagnostic information

Diagnosis of PCD has evolved over the years.[31] Therefore, CH-PCD includes patients diagnosed in different ways, depending on the year when they were diagnosed. For most patients, the diagnosis is based on a combination of tests in accordance with current recommendations; whereas for some, it was based on strong clinical suspicion (*e.g*. Kartagener syndrome) and the full diagnostic algorithm had not been completed. Using the European Respiratory Society’s diagnostic guidelines, we defined definite PCD as a biallelic PCD-causing mutation or a hallmark defect identified by transmission electron microscopy (TEM) [32,33]. We defined PCD as probable if there was either an abnormal light or high frequency video microscopy finding; low (≤ 77 nL/min) nasal nitric oxide; non-hallmark defect identified by TEM; pathologic immunofluorescence finding; or suspicions from genetic findings for PCD. In the absence of test results indicating PCD diagnosis or an incomplete algorithm, we defined PCD as a clinical diagnosis if there was a strong clinical suspicion. We used the international consensus guidelines (BEAT-PCD TEM criteria) to categorise ciliary ultrastructural defects identified by TEM [33].

### Statistical analysis

We described study population characteristics and prevalence, frequency and characteristics of reported symptoms overall and separately in children (<18 years) and adults, using median and interquartile range (IQR) for continuous variables and numbers and proportions for categorical variables. We compared prevalence and frequency of symptoms between males and females and by age using chi-square and t-tests. In a subsample of patients with abnormal TEM findings, we described and compared symptom frequency by ciliary ultrastructural defect. We performed all analyses using Stata version 16 (StataCorp LLC, Texas, USA).

## RESULTS

We had an excellent response rate with 74 out of 86 (86%) invited persons returning the questionnaire (Figure S1). The median age at survey was 23 years (IQR: 15.2–50-6) with 50 adults and 24 adolescents and children participating (Table 1). Most participants lived in German-speaking Swiss cantons (79%) and 38 (51%) were female.

**Table 1:** Characteristics of Swiss children and adults with PCD participating in the survey (N=74)

|  | **Total N (%)** | **Children <18 y N (%)** | **Adults ≥18 y N (%)** |
| --- | --- | --- | --- |
| **Number of participants** | **74 (100)** | 24 (100) | 50 (100) |
| **Age** | 23.2 (15.2-50.6) | 12.6 (5.9-15.0) | 32.4(23.1-57.0) |
| **Sex, female** | 38 (51) | 7 (29) | 31 (62) |
| **Area of residence** |  |  |  |
| German-speaking Switzerland | 58 (79) | 17 (71) | 41 (82) |
| French-speaking Switzerland | 16 (21) | 7 (29) | 9 (18) |
| **Diagnostic certainty** |  |  |  |
| Definite PCD diagnosis # | 38 (51) | 13(54) | 25 (50) |
| Probable PCD diagnosis ¶ | 23 (31) | 9(38) | 14 (28) |
| Clinical diagnosis only | 13 (18) | 2 (8) | 11 (22) |
| **Laterality defect** |  |  |  |
| Situs solitus | 38 (51) | 14 (58) | 24 (48) |
| Situs ambiguous | 2 (3) | 0 (0) | 2 (4) |
| Situs inversus | 32 (43) | 9 (38) | 23 (46) |
| Unknown | 2 (3) | 1 (4) | 1 (2) |
| **Congenital heart defect** | 5 (9) | 3 (13) | 2 (4) |
| **Retinitis pigmentosa** | 1 (1) | 0 (0) | 1 (2) |
| **Hydrocephalus** | 1 (1) | 0 (0) | 1 (2) |
| **Smoking** |  |  |  |
| Active smoking | 7 (9) | 0 (0) | 7 (14) |
| Ex-smoker | 4 (5) | 0 (0) | 4 (8) |
| Passive smoking (in household) | 18 (24) | 5 (21) | 13 (26) |
| **Occupation/ school** |  |  |  |
| Student/preschool | 29 (39) | 24 (100) | 5 (10) |
| 81-100% employment | 21 (28) | - | 21 (42) |
| 61-80% employment | 2 (3) | - | 2 (4) |
| 41-60% employment | 5 (9) | - | 5 (10) |
| ≤40% employment | 8 (11) | - | 8 (16) |
| Retired/ Disability pension | 9 (12) | - | 9 (18) |

PCD: primary ciliary dyskinesia, y: years. All characteristics are presented as N and column % except for age, which is presented as median and IQR: interquartile range.
#: biallelic pathogenic mutation or hallmark defect identified by electron microscopy
¶: at least one of the following: abnormal light or high frequency video microscopy finding/ low (≤77 nL/min) nasal nitric oxide value/non-hallmark defect identified by electron microscopy/pathologic immunofluorescence finding/ genetic findings suspicious for PCD

### PCD diagnosis

Half of participants had a definite PCD diagnosis based on current guidelines and 31% had a probable diagnosis with one or more tests indicating probable PCD. Among the remaining 13 patients (18%), PCD was diagnosed clinically after excluding other diagnoses; however, these patients had not undergone a complete diagnostic algorithm; and 9 of the 13 participants had typical Kartagener syndrome. We include more information about diagnostic test availability and results for the study population in Table S1.

### Participant characteristics

Thirty-two patients reported situs inversus and two other laterality defects. Five patients reported congenital heart defects; retinitis pigmentosa and hydrocephalus were only reported by one patient each. No patient reported renal problems. Seven adult patients reported occasional or frequent tobacco smoking, while an additional four participants were ex-smokers. Passive exposure to tobacco smoke in households was common; it was reported by 18 participants (24%). Age and sex did not differ between questionnaire respondents and non-respondents (data not shown). Missing responses to individual symptoms questions were less than 5%.

### Upper respiratory symptoms

Among upper respiratory symptoms, chronic nasal symptoms were most reported (Table 2 and Figure 1) and by 94% of participants. Most complained about a runny (65%) or blocked (55%) nose. Forty-six (62%) people said their nasal symptoms persisted all the time. More than one-third of participants reported anosmia or hyposmia during the past three months. Despite the high prevalence of nasal problems, only 9% reported frequent snoring; most said they snored only rarely or sometimes (53%) or never (38%). Headaches were reported by 69% of participants; and among 45% of participants even outside of infection. Only 9% reported headaches worsened when bending down—a typical sign of sinus infection. Ear pain was reported by more than half of participants; ear discharge was less common (19%). Children and adults reported hearing problems (58%), which were frequent among 22% of participants.

**Table 2:** Upper respiratory and ear symptoms of Swiss children and adults with PCD (N=74)

|  | **Total N (%)** | **Children <18 y N (%)** | **Adults ≥18 y N (%)** |
| --- | --- | --- | --- |
| **Number of participants** | 74 (100) | 24 (100) | 50 (100) |
| **Nasal symptoms** |  |  |  |
| Daily/often | 55 (74) | 15 (63) | 40 (80) |
| Sometimes/rarely | 15 (20) | 7 (29) | 8 (16) |
| Never | 4 (6) | 2 (8) | 2 (4) |
| **Nasal symptoms persist all the time** | 46 (62) | 13 (54) | 33 (66) |
| **Runny nose** | 48 (65) | 16 (67) | 32 (64) |
| **Blocked nose** | 41 (55) | 15 (63) | 26 (52) |
| **Sneezing** | 18 (24) | 3 (4) | 15 (30) |
| **Anosmia/hyposmia** | 28 (38) | 4 (17) | 24 (48) |
| **Snoring** |  |  |  |
| Daily/often | 7 (9) | 2 (8) | 5 (10) |
| Sometimes/rarely | 39 (53) | 13 (54) | 26 (52) |
| Never/not reported | 28 (38) | 9 (38) | 19 (38) |
| **Snoring also without having a cold/infection** | 23 (31) | 6 (25) | 17 (34) |
| **Snoring almost every night** | 9 (12) | 4 (17) | 5 (10) |
| **Headaches** |  |  |  |
| Daily/often | 12 (16) | 5 (21) | 7 (14) |
| Sometimes/rarely | 39 (53) | 9 (38) | 30 (60) |
| Never/not reported | 23 (31) | 10 (42) | 13 (26) |
| **Headaches also without having a cold/infection** | 33 (45) | 11 (46) | 22 (44) |
| **Headaches worst in the evening** | 30 (41) | 12 (50) | 18 (36) |
| **Headaches worst while bending down** | 7 (9) | 0 (0) | 7 (14) |
| **Ear pain** |  |  |  |
| Daily/often | 11 (15) | 4 (17) | 7 (14) |
| Sometimes/rarely | 32 (43) | 5 (21) | 27 (54) |
| Never/not reported | 31 (42) | 15 (62) | 16 (32) |
| **Ear discharge** |  |  |  |
| Daily/often | 3 (4) | 1 (4) | 2 (4) |
| Sometimes/rarely | 11 (15) | 2 (8) | 9 (18) |
| Never/not reported | 60 (81) | 21 (88) | 39 (78) |
| **Hearing problems** |  |  |  |
| Daily/often | 16 (22) | 3 (13) | 13 (26) |
| Sometimes/rarely | 27 (36) | 7 (29) | 20 (40) |
| Never/not reported | 31 (42) | 14 (58) | 17 (34) |

PCD: primary ciliary dyskinesia, y: years. All symptoms are presented as N and column %

**Figure 1.**
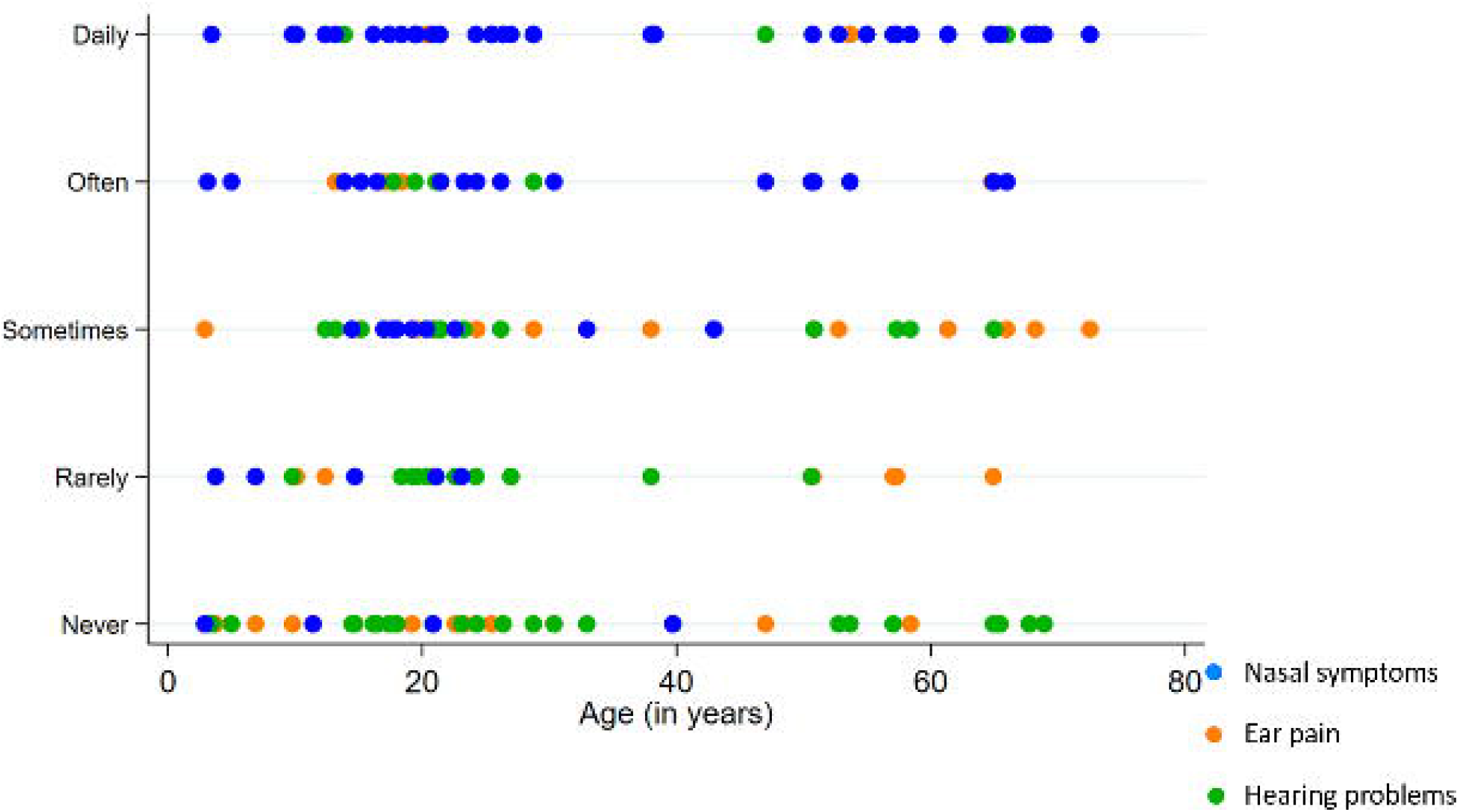
Prevalence and frequency of upper respiratory symptoms by age group among Swiss participants with PCD (N=74)

### Lower respiratory symptoms

Among lower respiratory symptoms, cough was reported by almost all participants (99%), and frequently by 72% (Table 3 and Figure 2). Twenty-nine (39%) respondents reported complications from excessive cough, most commonly gastroesophageal reflux (15%), vomiting (11%), and urinary incontinence (8%). Nearly all (99%) also reported excessive sputum, yet only 57% said they expectorated it. Wheezing was reported by 62% of patients and frequently by 12%; the most common wheezing triggers were infection and exercise (31%), followed by cold air and weather changes (12%), and house dust and intense laughter or crying (9%). Shortness of breath was reported by two-thirds of participants and frequently by 20%; respondents reported that shortness of breath was triggered mainly by exercising (51%) or walking up the stairs. Shortness of breath during normal daily activities or rest was reported by 9%. Almost half of adult participants reported normal dyspnea mMRC scores (0: I have shortness of breath with strenuous exercise only); 2 patients reported a high score (3: I stop for breath after walking about 100m or after a few minutes on level ground); and no respondents reported the highest score (4: I am too breathless to leave the house). Chest pain was reported by 54% and usually only as sometimes or rarely. The most common triggers were exercise and infection and 20% reported chest pain during normal daily activities or rest.

**Table 3:**
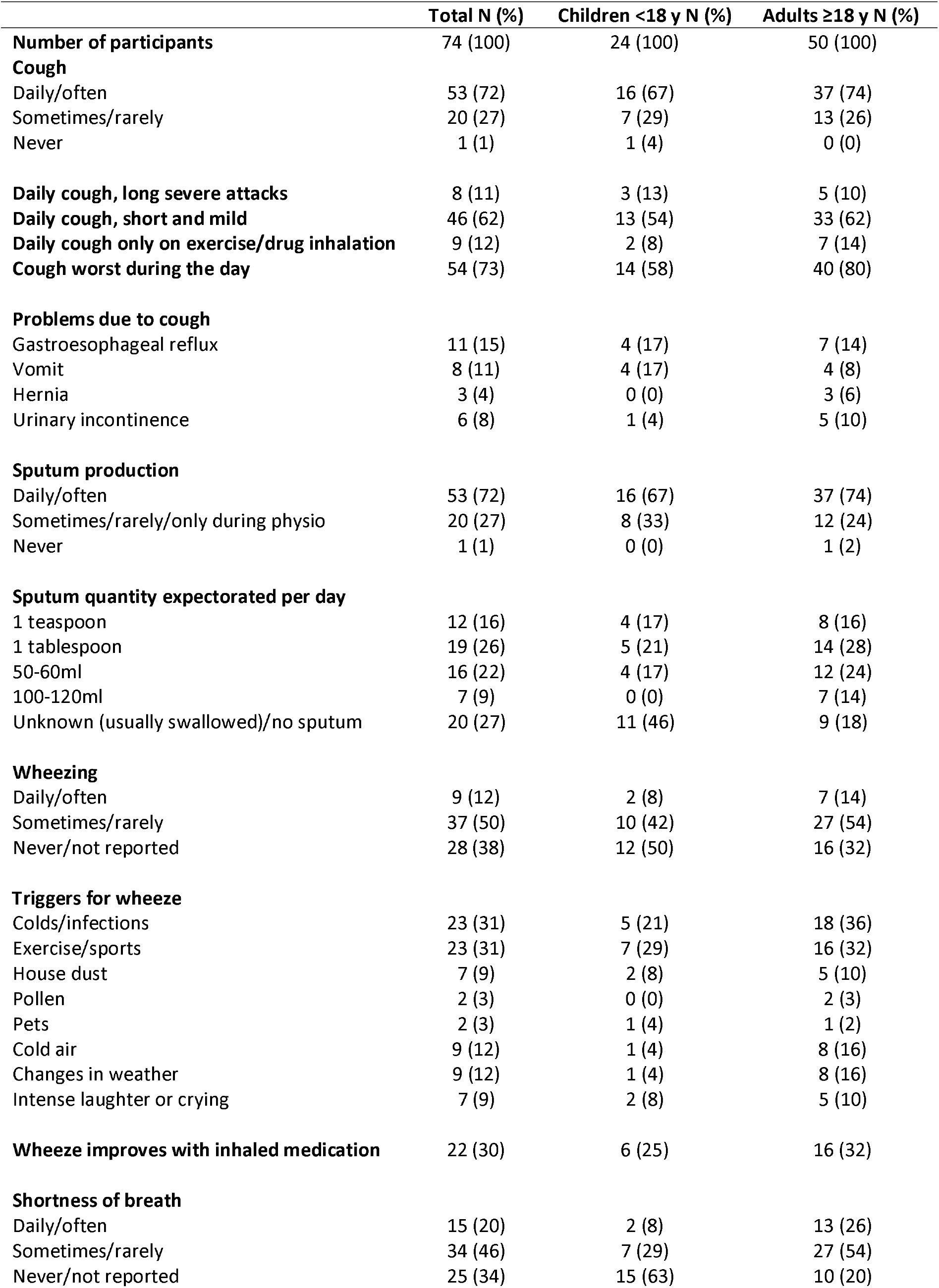
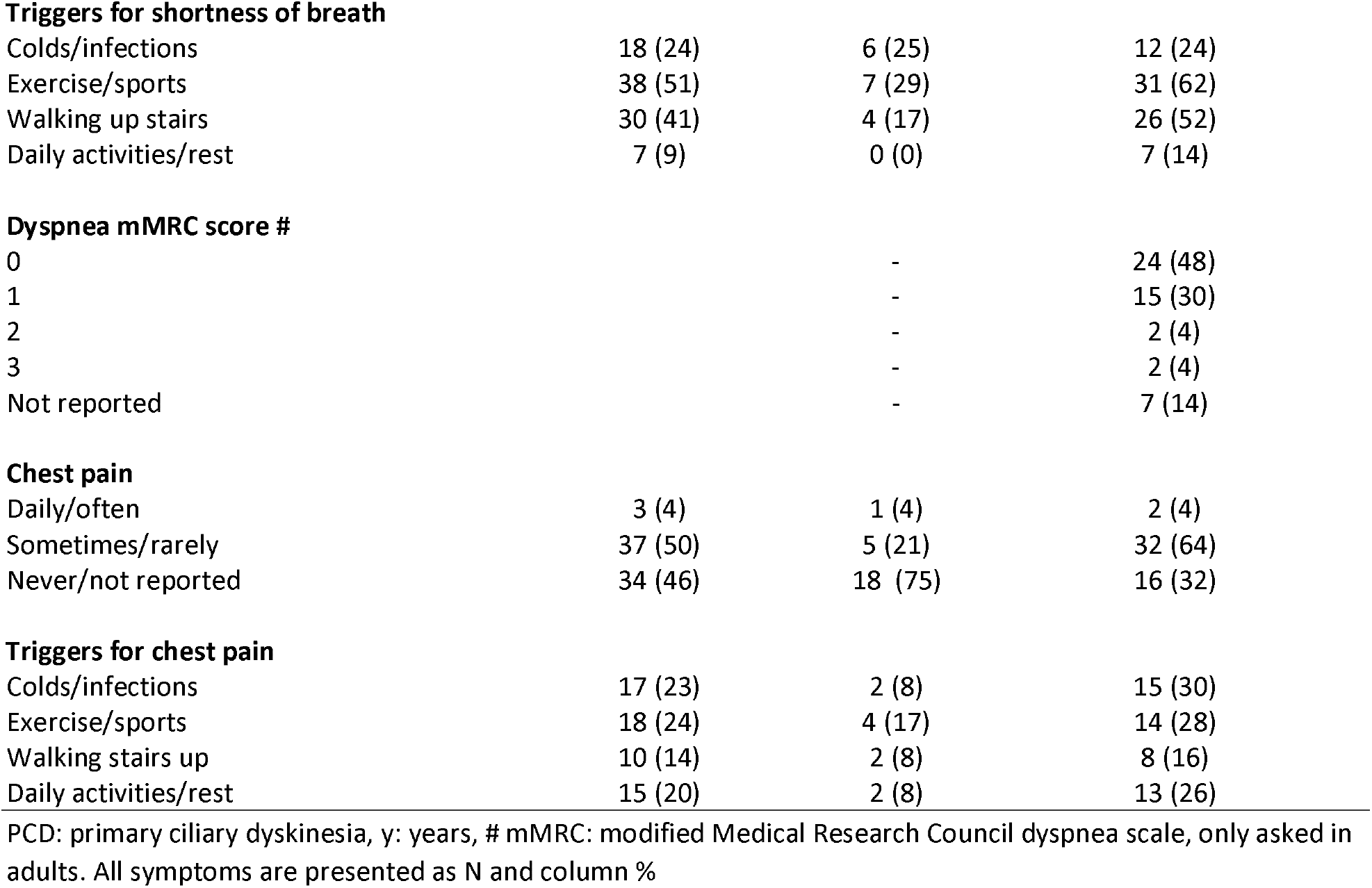
Lower respiratory symptoms of Swiss children and adults with PCD (N=74)

|  | **Total N (%)** | **Children <18 y N (%)** | **Adults ≥18 y N (%)** |
| --- | --- | --- | --- |
| **Number of participants** | 74 (100) | 24 (100) | 50 (100) |
| **Cough** |  |  |  |
| Daily/often | 53 (72) | 16 (67) | 37 (74) |
| Sometimes/rarely | 20 (27) | 7 (29) | 13 (26) |
| Never | 1 (1) | 1 (4) | 0 (0) |
| **Daily cough, long severe attacks** | 8 (11) | 3 (13) | 5 (10) |
| **Daily cough, short and mild** | 46 (62) | 13 (54) | 33 (62) |
| **Daily cough only on exercise/drug inhalation** | 9 (12) | 2 (8) | 7 (14) |
| **Cough worst during the day** | 54 (73) | 14 (58) | 40 (80) |
| **Problems due to cough** |  |  |  |
| Gastroesophageal reflux | 11 (15) | 4 (17) | 7 (14) |
| Vomit | 8 (11) | 4 (17) | 4 (8) |
| Hernia | 3 (4) | 0 (0) | 3 (6) |
| Urinary incontinence | 6 (8) | 1 (4) | 5 (10) |
| **Sputum production** |  |  |  |
| Daily/often | 53 (72) | 16 (67) | 37 (74) |
| Sometimes/rarely/only during physio | 20 (27) | 8 (33) | 12 (24) |
| Never | 1 (1) | 0 (0) | 1 (2) |
| **Sputum quantity expectorated per day** |  |  |  |
| 1 teaspoon | 12 (16) | 4 (17) | 8 (16) |
| 1 tablespoon | 19 (26) | 5 (21) | 14 (28) |
| 50-60ml | 16 (22) | 4 (17) | 12 (24) |
| 100-120ml | 7 (9) | 0 (0) | 7 (14) |
| Unknown (usually swallowed)/no sputum | 20 (27) | 11 (46) | 9 (18) |
| **Wheezing** |  |  |  |
| Daily/often | 9 (12) | 2 (8) | 7 (14) |
| Sometimes/rarely | 37 (50) | 10 (42) | 27 (54) |
| Never/not reported | 28 (38) | 12 (50) | 16 (32) |
| **Triggers for wheeze** |  |  |  |
| Colds/infections | 23 (31) | 5 (21) | 18 (36) |
| Exercise/sports | 23 (31) | 7 (29) | 16 (32) |
| House dust | 7 (9) | 2 (8) | 5 (10) |
| Pollen | 2 (3) | 0 (0) | 2 (3) |
| Pets | 2 (3) | 1 (4) | 1 (2) |
| Cold air | 9 (12) | 1 (4) | 8 (16) |
| Changes in weather | 9 (12) | 1 (4) | 8 (16) |
| Intense laughter or crying | 7 (9) | 2 (8) | 5 (10) |
| **Wheeze improves with inhaled medication** | 22 (30) | 6 (25) | 16 (32) |
| **Shortness of breath** |  |  |  |
| Daily/often | 15 (20) | 2 (8) | 13 (26) |
| Sometimes/rarely | 34 (46) | 7 (29) | 27 (54) |
| Never/not reported | 25 (34) | 15 (63) | 10 (20) |
| **Triggers for shortness of breath** |  |  |  |
| Colds/infections | 18 (24) | 6 (25) | 12 (24) |
| Exercise/sports | 38 (51) | 7 (29) | 31 (62) |
| Walking up stairs | 30 (41) | 4 (17) | 26 (52) |
| Daily activities/rest | 7 (9) | 0 (0) | 7 (14) |
| **Dyspnea mMRC score #** |  |  |  |
| 0 |  | - | 24 (48) |
| 1 |  | - | 15 (30) |
| 2 |  | - | 2 (4) |
| 3 |  | - | 2 (4) |
| Not reported |  | - | 7 (14) |
| **Chest pain** |  |  |  |
| Daily/often | 3 (4) | 1 (4) | 2 (4) |
| Sometimes/rarely | 37 (50) | 5 (21) | 32 (64) |
| Never/not reported | 34 (46) | 18 (75) | 16 (32) |
| **Triggers for chest pain** |  |  |  |
| Colds/infections | 17 (23) | 2 (8) | 15 (30) |
| Exercise/sports | 18 (24) | 4 (17) | 14 (28) |
| Walking stairs up | 10 (14) | 2 (8) | 8 (16) |
| Daily activities/rest | 15 (20) | 2 (8) | 13 (26) |

PCD: primary ciliary dyskinesia, y: years, # mMRC: modified Medical Research Council dyspnea scale, only asked in adults. All symptoms are presented as N and column %

**Figure 2.**
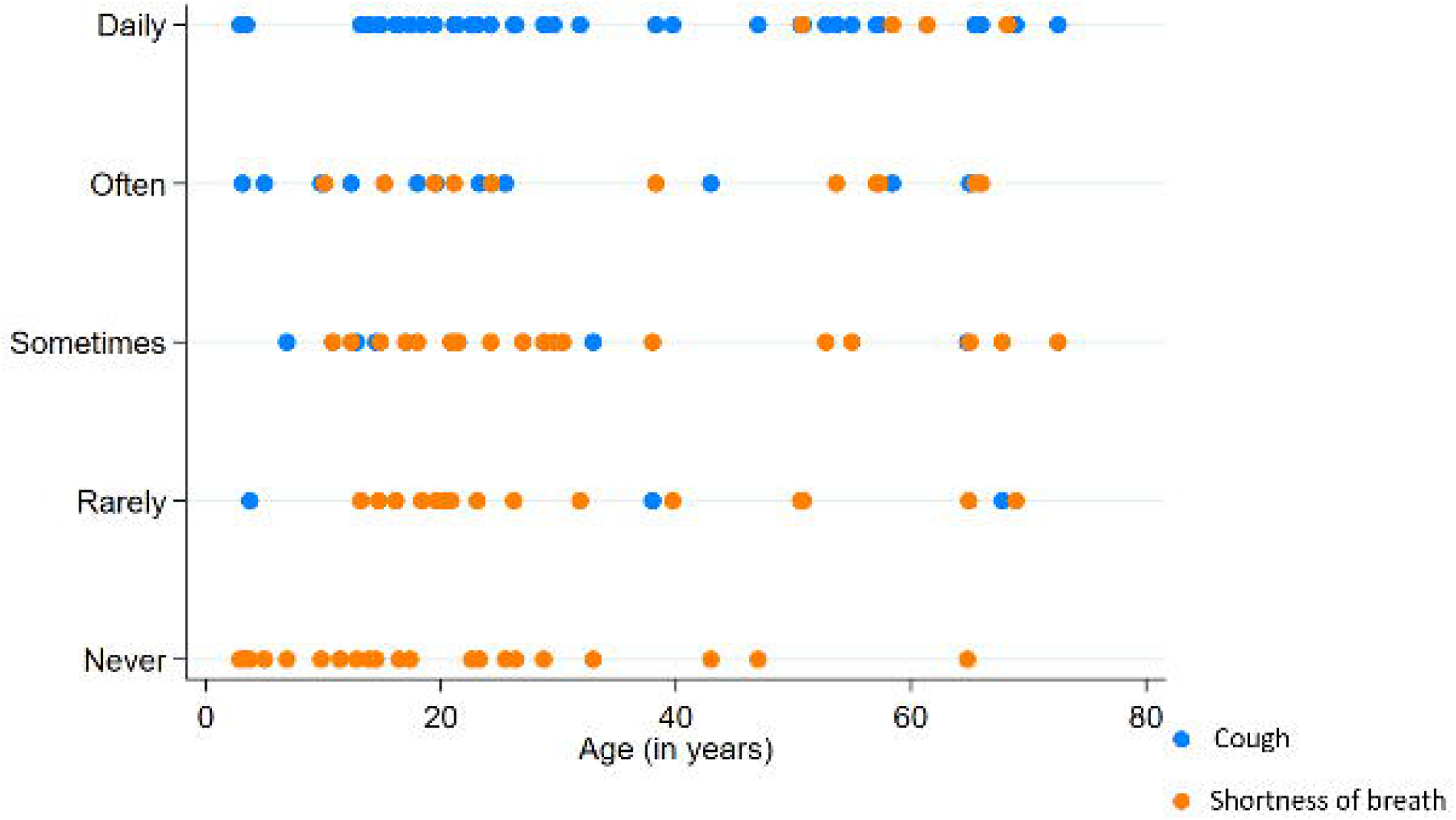
Prevalence and frequency of lower respiratory symptoms by age group among Swiss participants with PCD (N=74)

### Differences by age and sex

Patients who reported frequent nasal symptoms were older (mean, 95% CI: 34.5, 28.8–40.2 years) compared to patients who reported nasal symptoms less frequently or never (18.6, 13.19–23.9 years) (p= 0.001). Anosmia was reported more frequently by adult patients (p=0.009). Figure 3 displays the frequency of upper respiratory symptoms by age in the study population; it shows that all older patients reported nasal symptoms often or daily. For lower respiratory symptoms, patients who reported frequent shortness of breath were older (44.4, 32.9–55.9 years) compared to patients who reported it less frequently or never (26.9, 22.0– 31.9 years) (p=0.001). Figure 4 shows that younger patients rarely reported daily shortness of breath compared to older patients (p=0.006). Chest pain was also more commonly reported by adults (p=0.007). We found no difference in prevalence and frequency of respiratory symptoms by sex (Figures S2-S3).

**Figure 3.**
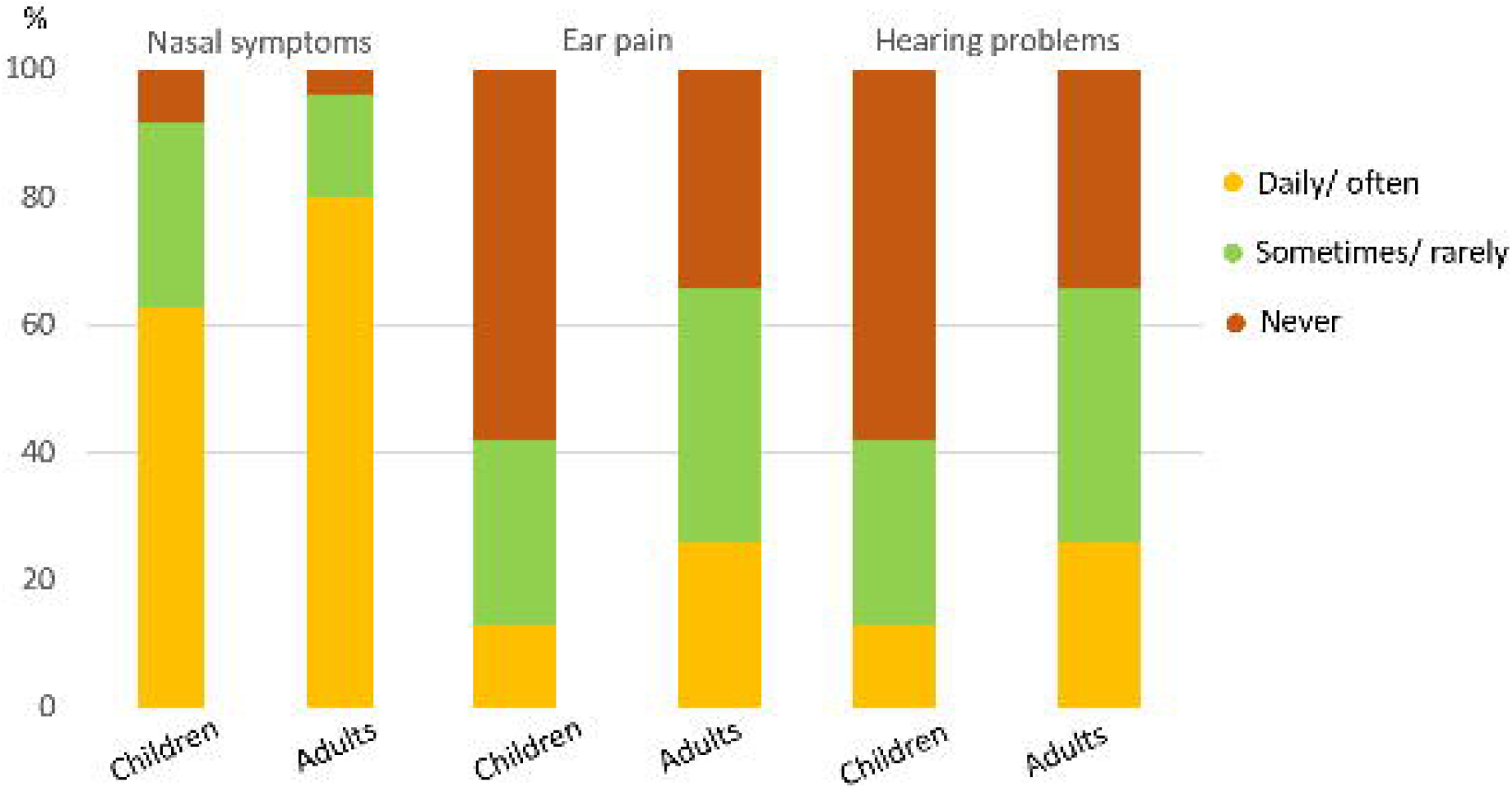
Frequency of upper respiratory symptoms by age (in years) among Swiss participants with PCD. Each colour represents a symptom and each dot represents one patient. The density of dots corresponds to how many people within the same age range reported a symptom of equal frequency (daily, often, sometimes, rarely, or never).

**Figure 4.**
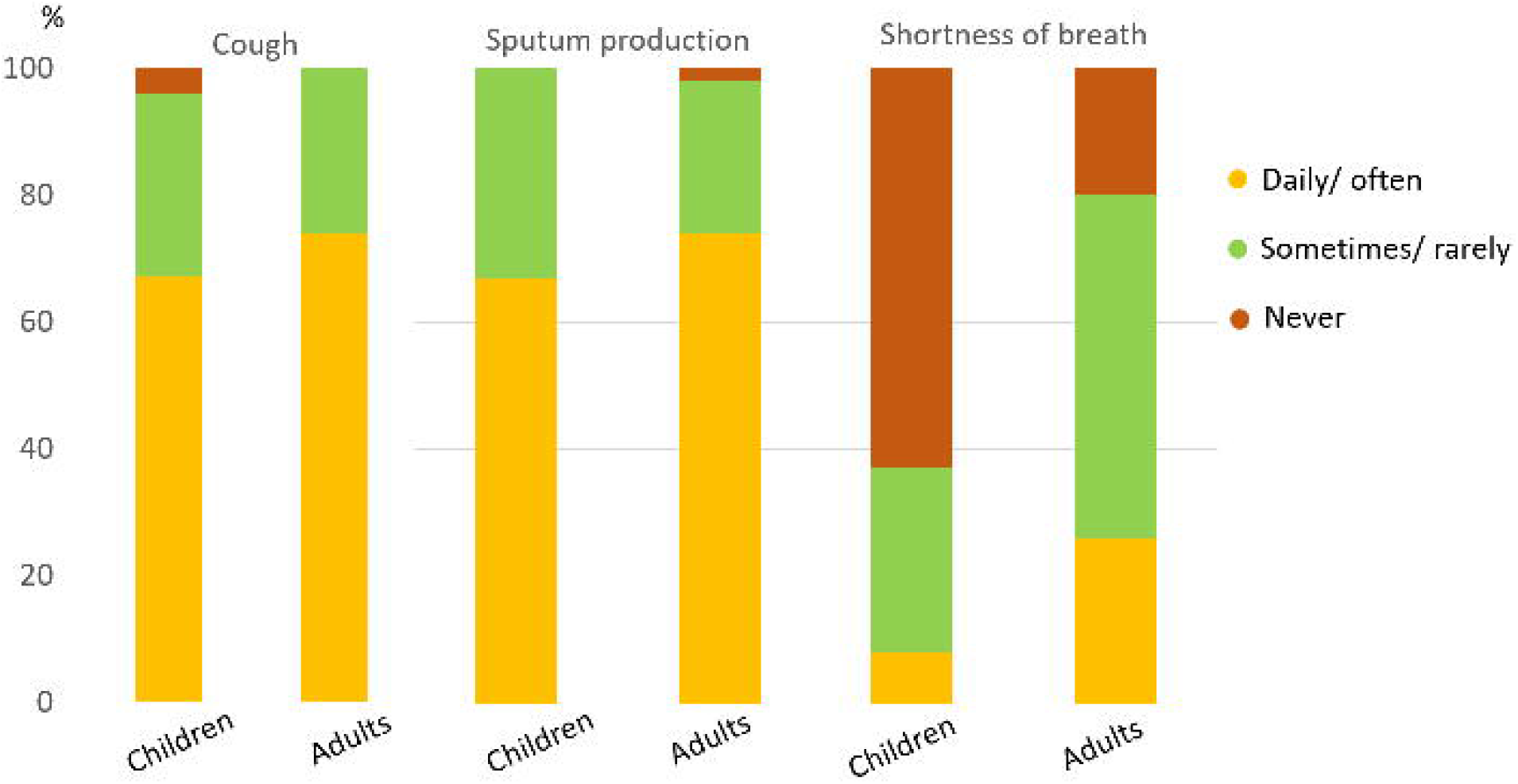
Frequency of lower respiratory symptoms by age (in years) among Swiss participants with PCD. Each colour represents a symptom and each dot represents one patient. The density of dots corresponds to how many people within the same age range reported a symptom of equal frequency (daily, often, sometimes, rarely or never).

### Differences by ultrastructural defect

Out of the 45 patients who had undergone EM analysis, 40 had a reported ultrastructural defect (Table S1). The most common were class 1 defect in outer and inner dynein arms (ODA and IDA, 18%) followed by isolated ODA defects (15%). We found no difference in prevalence and frequency of reported respiratory symptoms between different ultrastructural defect categories in this subgroup of 40 patients (Figure S4).

## DISCUSSION

Ours is the first study about patient-reported PCD-specific symptoms. We found people reported a variety of respiratory symptoms; the most common were nasal symptoms and cough and sputum production. Over half reported ear problems, particularly ear pain and hearing difficulties. One out of five reported frequent shortness of breath. Nasal problems and shortness of breath became more frequent with age; we found no differences between sexes.

A strength of our study is the use of the PCD-specific and standardised FOLLOW-PCD questionnaire [25], which led to homogeneous, standardised in-depth data about symptoms reported directly from participants or parents of participants. We also had a low proportion of missing data. Another strength was the representative study population and the high response rate. Since the study was nested in the national CH-PCD registry, it allowed for an unselected population of people with PCD, including children and adults [20].

### Limitations

Although the sample size is reasonable for a rare disease in a small country, it did not allow for detailed subgroup analyses for more than two age groups. Also, our cross-sectional survey represents a snapshot of the clinical status of Swiss patients with PCD and did not allow studying symptom variations by season. It is possible that our results were influenced by lifestyle and health care changes during the SARS-CoV-2 pandemic. However, over three-fourths of the questionnaires were returned by March 2020, so we suspect small effects.

### Comparison with other studies

Previous data about PCD clinical symptoms come mostly from nonstandard reporting found during chart reviews. Therefore, it is not easy to compare prior studies with our results. Furthermore, few questionnaire-based studies focus on quality of life or the psychosocial impact of the disease [34]. In fact, only one prior questionnaire survey assessed symptoms and the psychosocial impact of PCD among 93 people with PCD in the UK [35]. Through the UK PCD support group, the study took place 18 years ago and used the St. George’s Respiratory Questionnaire (SGRQ) [36]. The authors reported respiratory symptom scores declined after age 25. They also found that almost all participants reported “a runny nose and nasal congestion,” while “pain over the sinuses” and “headaches” were less frequent [35]. We did not find an increase of all symptoms with advancing age. However, direct comparisons are not feasible due to incommensurable differences between SGQR and the FOLLOW-PCD questionnaire [25]. In our study, 14% of adult participants smoked cigarettes daily or occasionally. Smoking prevalence was much lower than in the general Swiss population (23.5% for persons over 15 years) [37] but higher than previous reports in people with cystic fibrosis. [38]

Detailed standardised information about PCD symptoms is important for studying phenotypic variability of PCD. Recent studies suggest that PCD is not a mild disease. Most Swiss patients reported frequent nasal symptoms, cough, and sputum, and 20% reported shortness of breath as a frequent occurrence triggered by infections, exercise, and regular activities. We also showed that nasal symptoms deteriorate with age; all older adults report them frequently. Therefore since upper airway management remains important, regular multidisciplinary follow-up is also needed for adults. Our survey also highlighted how different patients experience respiratory symptoms and that there are patients who report mild symptoms only. This could partly be explained by the fact that people with PCD are accustomed to chronic symptoms, consider them normal and underreport these symptoms. Yet, it could also indicate distinct PCD phenotypes. Standardising detailed clinical information about PCD symptoms and using appropriate methodological approaches in large collaborative studies are needed to better characterise PCD’s phenotypic variability and describe phenotypic groups [23,39-41].

For the first time, a study provides patient-reported data about a wide range of PCD-specific symptoms. Standardised data collected throughout life with repeated surveys will allow studying disease course and describing associations between phenotypes and structural and functional respiratory changes. A better understanding of the variability of the clinical picture can inform management and support individualised treatment approaches in the future.

## Data Availability

All data produced in the present study are available upon reasonable request to the authors

## Author Contributions

M Goutaki and CE Kuehni developed the concept and designed the study. M Goutaki, L Hüsler, and YT Lam organised the survey, then cleaned and standardised the data. M Goutaki performed the statistical analyses and drafted the manuscript. All authors commented and revised the manuscript. M Goutaki and CE Kuehni take final responsibility for content.

## Acknowledgments

We want to thank all the people with PCD in Switzerland and their families for participating in the survey and the CH-PCD. We are also grateful to the Swiss PCD support group that closely collaborates with us. We thank Eugenie Collaud (Institute of Social and Preventive Medicine [ISPM], University of Bern) for her contributions to the French translation of the questionnaire and Kristin Marie Bivens (ISPM, University of Bern) for her editorial assistance.

The current Swiss PCD research group includes (in alphabetical order): Juerg Barben (Kantonsspital St. Gallen), Sylvain Blanchon (University Hospital of Lausanne), Jean-Louis Blouin (University Hospitals of Geneva), Marina Bullo (Inselspital Bern), Carmen Casaulta (Inselspital Bern), Cristian Clarenbach (University Hospital of Zurich), Myrofora Goutaki (ISPM, University of Bern), Nicolas Gürtler (University Hospital of Basel, Switzerland), Beat Haenni (Institute of Anatomy Bern), Andreas Hector (University Children’s Hospital Zurich), Michael Hitzler (Children’s Hospital Lucerne), Andreas Jung (University Children’s Hospital Zurich), Lilian Junker (Hospital Thun), Elisabeth Kieninger (Inselspital Bern), Claudia E. Kuehni (ISPM, University of Bern), Yin Ting Lam (ISPM, University of Bern), Philipp Latzin (Inselspital Bern), Romain Lazor (University Hospital of Lausanne), Dagmar Lin (Bern University Hospital), Marco Lurà (Children’s Hospital Lucerne), Loretta Müller (Inselspital Bern), Eva Pedersen (ISPM, University of Bern), Nicolas Regamey (Children’s Hospital Lucerne), Isabelle Rochat (University Hospital of Lausanne), Daniel Schilter (Quartier Bleu Bern), Iris Schmid (Quartier Bleu Bern), Bernhard Schwizer (Quartier Bleu Bern), Andrea Stokes (Inselspital Bern), Daniel Trachsel (University Children’s Hospital Basel), Stefan A. Tschanz (Institute of Anatomy, University of Bern), Johannes Wildhaber (University of Fribourg), and Maura Zanolari (Hospital of Bellinzona).

