## Supplementary material for "Respiratory symptoms of Swiss people with Primary Ciliary Dyskinesia": Figure S1

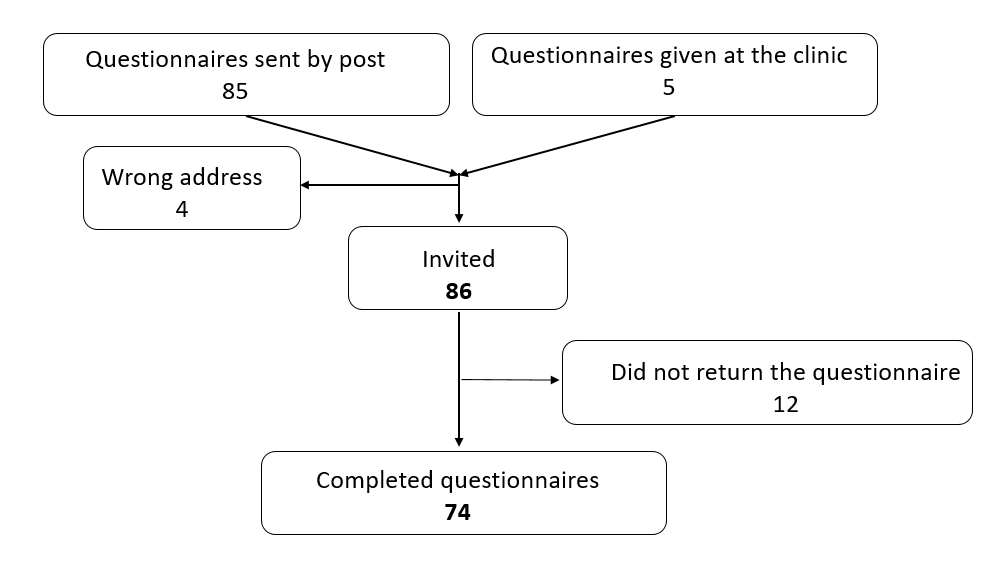

**Figure S1** Flow chart of Swiss patients with PCD or their parents who were invited and participated in the survey.

**Table S1:** Availability and results of diagnostic tests among Swiss patients (N=74) with PCD

|  | **N (%)** |
| --- | --- |
| **nNO testing** |  |
| nNO normal levels | 6 (8) |
| nNO low levels (≤77 nL/min)  Not performed | 37 (50)  31 (42) |
| **Videomicroscopy** |  |
| Beat frequency/pattern normal | 10 (14) |
| Beat frequency/pattern abnormal | 35 (47) |
| Not performed | 29 (39) |
| **Electron microscopy** |  |
| Normal ultrastructure | 5 (7) |
| Class 1 defect identified | 30 (41) |
| ODA | 11 (15) |
| ODA and IDA | 13 (18) |
| MTD and IDA | 6 (8) |
| Class 2 defect identified | 7 (9) |
| Inconclusive result | 3 (4) |
| Not performed | 29 (39) |
| **Immunofluorescence analysis** |  |
| Normal | 3 (4) |
| Abnormal | 11 (15) |
| Not performed | 60 (81) |
| **Genetic analysis** |  |
| No pathogenic mutation identified | 2 (3) |
| Confirmed pathogenic mutation# | 11 (15) |
| Mutation only in one allele | 7 (9) |
| Not performed | 54 (73) |

PCD: primary ciliary dyskinesia; nNO: nasal nitric oxide; ODA: outer dynein arm defect; ODA and IDA: outer and inner dynein arm defect; MTD and IDA: microtubular disorganization and inner dynein arm defect

\# mainly DNAH5, DNAH11, DNAI1, HYDIN, CCDC39 and CCDC40 mutations

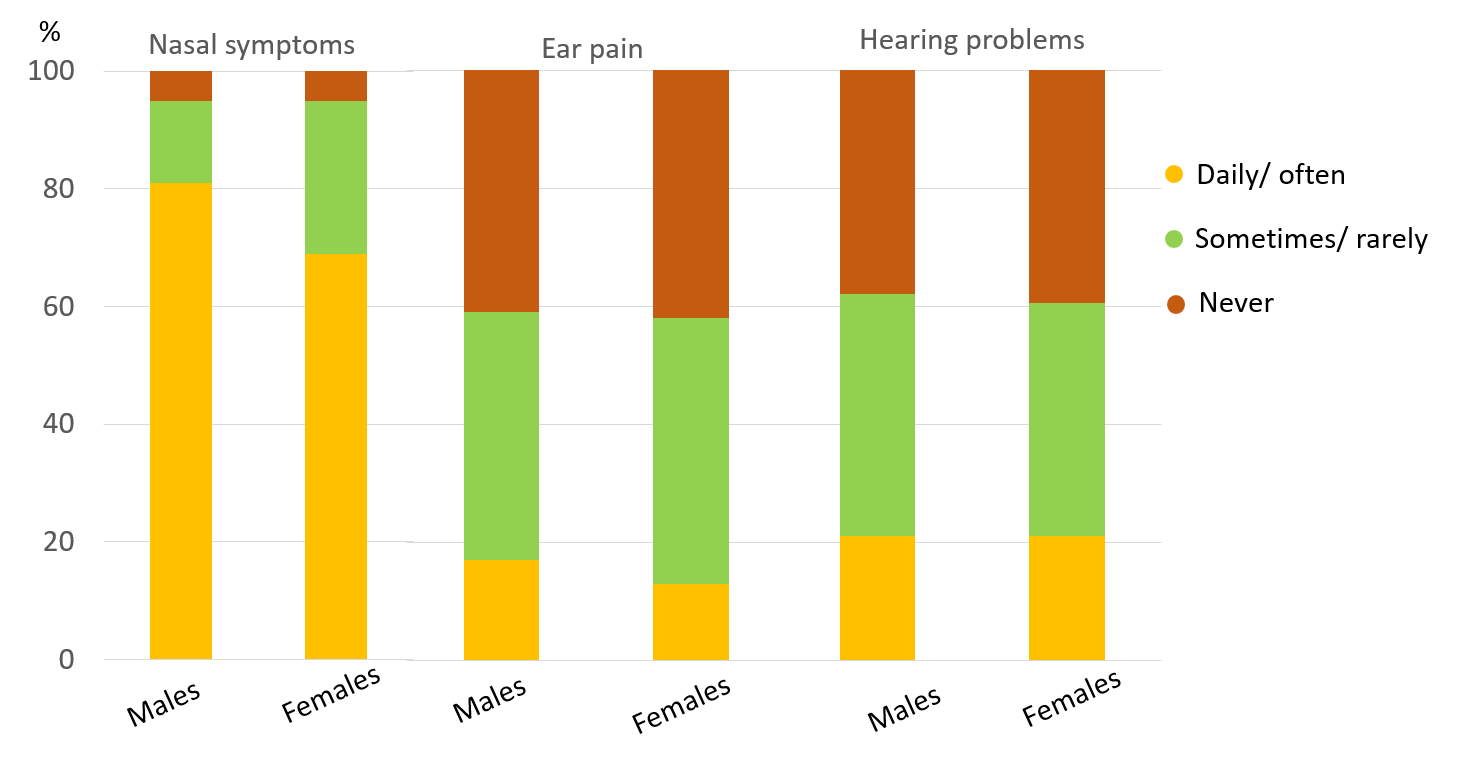

**Figure S2** Prevalence and frequency of upper respiratory symptoms by sex among Swiss participants with PCD

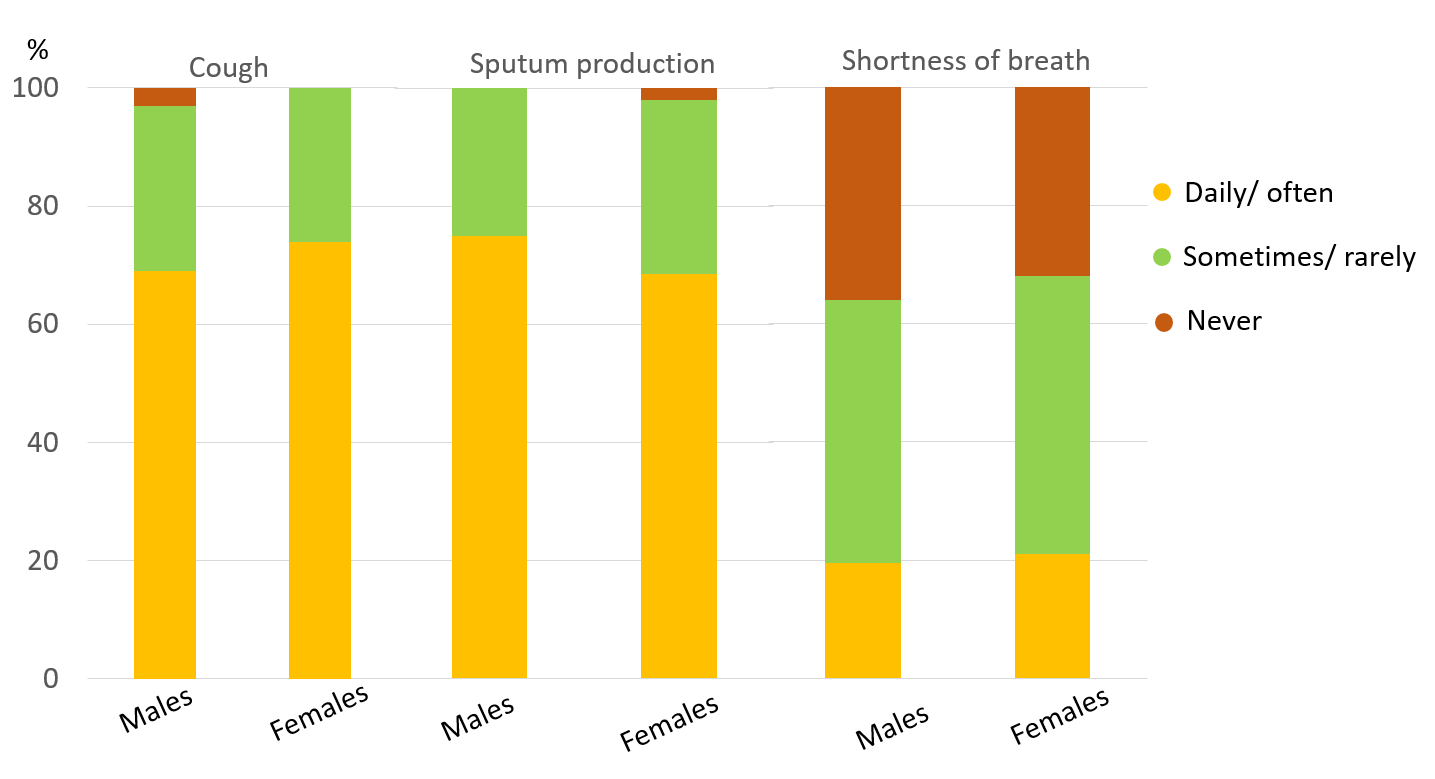

**Figure S3** Prevalence and frequency of lower respiratory symptoms by sex among Swiss participants with PCD

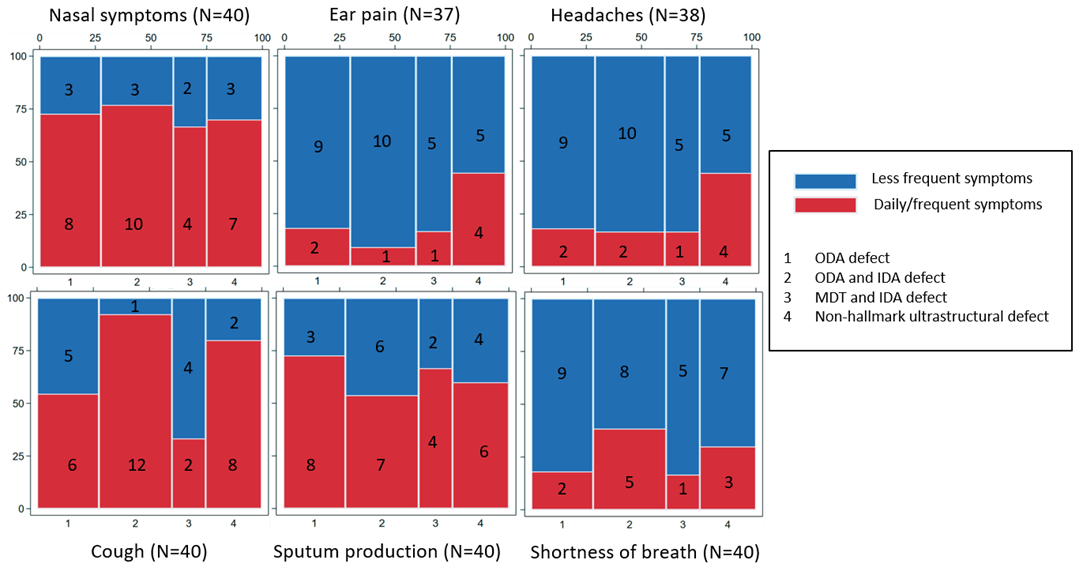

**Figure S4** Mosaic plot representing symptom frequency by ciliary ultrastructural defect among Swiss participants with PCD and with abnormal electron microscopy findings (N=40)

ODA: outer dynein arm defect; ODA and IDA: outer and inner dynein arm defect; MTD and IDA: microtubular disorganization and inner dynein arm defect
